# Oral health vulnerability and its associated risk factors among tribal elderly people in Bangladesh: a pilot study

**DOI:** 10.1101/2023.07.25.23293159

**Authors:** Shomrita Barua, Sunanda Bosu, Mohammad Farhadul Haque, Liza Bosak, Md Rezowan Rashid, Shaikh Riaduz Zaman, Md. Foyzur Rahman, Mohammad Meshbahur Rahman

## Abstract

**Background:** Tribal elderly is a vulnerable population due to their geographic location and lack of access to healthcare services. This study aims to assess the oral health status of tribal elderly in Bangladesh and identify any associated risk factors.

**Methods:** This cross-sectional pilot survey was conducted among 280 tribal elderlies aged 60 years and above residing in the main tribal residing region in Bangladesh. The data was collected through cluster sampling methods using a semi-structured questionnaire and oral examination. Different statistical tools including frequency distribution, Chi-square association test and multivariable logistic regression model were performed in data analysis.

**Results:** The results showed that 25.35% of the tribal elderly had high DMFT score, indicating poor oral health. Respondents’ age, sex, marital status, occupational status, sweet eater, tobacco, and alcohol consumption were significantly associated with oral health. The major oral clinical sign and symptoms were dental carries, teeth sensitivity, loose teeth, broken teeth, calculus and staining in teeth, tongue pain and bad breath. The logistic regression analysis suggested that elderly aged 80 and above had 3.33 times more, males were 4.6 time less, tobacco user (smoked/smokeless) were 2.03 times more and alcohol consumers were 6.83 times more likely to experienced poor oral health condition than their counterparts.

**Conclusions:** Elderly individuals were found to be more likely to suffer from poor oral health condition than younger individuals, and certain lifestyle factors such as tobacco and alcohol consumption, meal frequency, and sweet consumption can further increase the risk of poor oral health condition.

## Introduction

Oral disease is a prevalent cause of medical and emergency room visits that is closely linked to aging [1,2]. Poor oral health can negatively impact overall health and quality of life, leading to poor nutritional status among the elderly [3,4]. Consuming carbohydrates, sticky foods, and sweetened foods can contribute to poor oral health [5,6], as can the use of tobacco and alcohol [7,8]. Sweetened food and sticky food consumption leads to Dental caries, Gingival bleeding, periodontitis, calculus and other types of oral health problems [9–11]. Additionally, oral health problems can cause early loss of teeth and negatively impact the ability to chew food [3]. Geriatric individuals are particularly vulnerable to dental diseases due to lack of education and access to care [12–14], suffer from different types of dental diseases [15,16].

In South Asia, tribal elderly individuals often face significant oral health challenges. Studies have shown that the prevalence of dental caries among this population is high, with over 60% of individuals over the age of 60 experiencing the condition [17–19]. Additionally, periodontal disease is prevalent, with nearly 40% of tribal elderly individuals in the region affected [19–21]. These rates are significantly higher than those seen in non-tribal populations in South Asia. Factors such as limited access to dental care, poor oral hygiene practices, and a lack of education about oral health contribute to these high rates of oral health issues among tribal elderly individuals in the region [22,23].

Bangladesh, one of the most densely-populated countries in the world where 1.10% of the country’s total population are tribal [24]. They are mainly live in the flatland districts of the north and southeast of the country, while the rest reside in the Chittagong Hill Tracts [25]. Most of the people of tribes lives in very deep of the jungle and the hills. Smokeless tobacco has been implicated as a risk factor for numerous oral conditions, starting from gingivitis to oral cancers [26,27]. Many geriatric people among the tribes have the habit of chewing betel nuts, tobacco leaf without knowing its side effect [25,28].

Research on the oral health of tribal elderly individuals in Bangladesh has shown that they have a higher incidence of oral health issues compared to the general population [29–31]. One study found that almost 75% of tribal elderly individuals in Bangladesh had at least one decayed or missing tooth, and over 50% had periodontal disease [32,33]. This contrasts with the national average, where only about 60% of older adults have at least one decayed or missing tooth and around 25% have periodontal disease [32,34,35]. Factors that contribute to the poor oral health of tribal elderly individuals in Bangladesh include limited access to dental care, poverty, and poor oral hygiene practices. Additionally, cultural and traditional practices, such as chewing betel nut and tobacco, can also increase the risk of oral health issues [17,23,36]. Despite these challenges, there are programs and initiatives in place to improve oral health and access to dental care for tribal elderly individuals in Bangladesh.

The isolation from mainstream development activities, together with poverty and difficult healthcare accessibility made the tribal communities specifically vulnerable to various problems including oral health [25,37]. But relatively few studies have focused specifically on the tribal groups in the country [38,39]. Research on tribal health has predominantly focused on the prevalence of good oral health, profiles of dental illnesses, and health-provision coverage rather than people’s knowledge, practices, opinions of and attitudes towards health provision in the tribal areas. Therefore, this study aims to assess the oral health status of tribal elderly in Bangladesh and identify the associated risk factors.

## Methods

### Ethical consent and permission for data collection

This study followed the guidelines of World Medical Association (WMA) Declaration of Helsinki. The ethical approval was taken the institutional review board of the National Institute of Preventive and Social Medicine, Dhaka (IRB registration number: NIPSOM/IRB/2019/111) and formal permission of data collection in the community was taken from the tribal community leaders (called ‘*Karbari*’). Both written and verbal consent was taken from each participant before initiating the interview for data collection. A brief introduction on the aims and objectives of the study was given first and then, the written consent translated in native language was read out for illiterate tribal elderly. Participants who were agreed with the consent were finally included in the study.

### Study design

As a pilot initiative, this study was conducted cross-sectionally involving a person-centered general health assessment and a self-administered questionnaire in this study.

### Sample size, participants, and data collection

In this study, the estimated sample size was found 255 using the proportional sample size calculation formula at 5% margin of error. Considering an additional 10% non-response rate, the final sample size became 280 elderlies residing in tribal community in Bangladesh.

The study was conducted among elderlies aged 60 years and above in South-Eastern Bangladesh from June 2020 to May 2021. Simple random sampling (SRS) technique was used to collect the data. The residence of the tribal elderlies was divided into 20 *paras* (Para, social geographical zone, or village in Bangladesh). Eight *paras* were randomly selected by SRS, and then the eligible elderlies from each *para* were included for further interview. Face-to-face interview technique was used to collect socio-demographic data along with instruments for anthropometric data. The data were collected by a trained data enumerator (dental students) using the necessary data collection tools-Caries probe, Periodontal Probe and Dental Mirror. The inclusion criteria for data collection were: (i) aged 60 and above; (ii) living in the tribal community of South-Eastern Bangladesh; (iii) agreed to participate in a general dental health assessment and an interview.

### Variable measurements

Socio-demographic variables of the tribal elderly were age (in complete year), sex, religion, marital status, monthly family income, educational status, occupation and family type. Personal habit related variables were tobacco consumption and alcohol consumption. Diet related variables were meal time, variety of food taking, daily frequency of meal (dinner, lunch, breakfast), snacks eating, oil, drinking of tea, soft-drinks, sweet and vegetable consumption. The dietary habit was measured by 24 hours recall methods and 7 days food frequency method and other food related questions.

The patients were asked about their oral health conditions. A modified scoring system of WHO was used to detect gingival and periodontal status among the patients [40]. This score system was used to determine the periodontal health and gingival condition. They were asked to open their mouth and after checking oral conditions based upon clinical sign symptoms and asking questions the answers were noted down on the questionnaire. Dependent variable of the study was oral health status assessed by observing oral health related indicators, and Decayed, Missing due to caries, and Filled Teeth (DMFT). The DMFT score was computed for each elderly following the guideline of World Health Organization (WHO) oral health survey [41].

### Statistical analysis

The collected data were processed, edited, and coded first, and observed consistency by frequency distribution. In data analysis, descriptive statistics including frequency distribution table, graphs and diagrams were performed first. Elderly oral health status was assessed by DMFT levels. For analytical purpose, the DMFT index score was statistically classified into high DMFT (poor oral health status; DMFT score>5.0) and low DMFT (good oral health status; DMFT score<5.0) considering 75% percentiles (75% percentiles of DMFT was 5.0) which also supports WHO guideline on oral health survey [41]. The Pearson Chi-square test performed to observe the significantly associated factors of DMFT. The degree of associated factors of DMFT as well as oral health status was assessed by adjusted odds ratio in multivariable logistic regression model [12,42]. To perform multivariable logistic regression model, we converted our dependent variable DMFT as dichotomous [poor oral health status (high DMFT) coded as 1 and good oral health status (low DMFT) coded as 0]. Dataset management and all the statistical analysis was carried out through IBM SPSS Statistics 26.0. The confidence interval was used in 95% and level of confidence was set at 0.05.

## Results

### Background characteristics of the tribal elderly

The second column of Table 1 represents the socio-demographic characteristics of the tribal elderly. Among 280 respondents, most (76.8%) of the tribal elderly were 60-69 years of old following 14.3% were 70-79 and 8.9% were in the age group 80 and above. The proportion of female elderly was higher (63.3% vs. 36.8%) than male and majority (72.1%) of them were Buddhist in religion. Majority of the tribal elderly are poor and live in lower socio-economic condition where agriculture, day laborer and housewife were the main occupational status. Illiteracy rate was high among the tribal community. One-fifth of the tribal elderly live in joint and extended family and almost 50% of the family’s monthly income was only 10001-20000 Taka ($100-$200) (Table 1).

**Table 1.** Socio-demographic characteristics as the associated factors of DMFT.

| Characteristics | Frequency<br>n(%) | Low<br>DMFT (<5)<br>n(%) | High DMFT<br>(≥5)<br>n(%) | P# value |
| --- | --- | --- | --- | --- |
| **Age** |  |  |  |  |
| 60-69 years | 215(76.8) | 170(79.1) | 45(20.9) | 0.007*** |
| 70 to 79 years | 40(14.3) | 25(62.5) | 15(37.5) |  |
| >80 Years | 25(8.9) | 14(56.0) | 11(44.0) |  |
| **Sex** |  |  |  |  |
| Female | 177(63.2) | 135(76.3) | 42(23.7) | 0.08* |
| Male | 103(36.8) | 74(71.8) | 29(28.2) |  |
| **Marital Status** |  |  |  |  |
| Married | 228(81.4) | 176(77.2) | 52(22.8) | 0.04** |
| Unmarried/widowed/divorce | 52(18.6) | 33(63.5) | 19(36.5) |  |
| **Religion** |  |  |  |  |
| Buddhism | 202(72.1) | 154(76.2) | 48(23.8) | 0.32 |
| Christian and others | 78(27.9) | 55(70.5) | 23(29.5) |  |
| **Educational status** |  |  |  |  |
| Literate | 62(22.1) | 43(69.4) | 19(30.6) | 0.27 |
| Illiterate | 218(77.9) | 166(76.1) | 52(23.9) |  |
| **Occupational status** |  |  |  |  |
| Retire and dependent | 25(8.9) | 14(56.0) | 11(44.0) | 0.014*** |
| Housewife | 98(35.0) | 73(74.5) | 25(25.5) |  |
| Agriculture/Labour/Other | 107(38.2) | 89(83.2) | 18(16.8) |  |
| Business/Employed | 50(17.9) | 33(66.0) | 17(34.0) |  |
| **Family Type** |  |  |  |  |
| Single and Nuclear Family | 46(16.4) | 36(78.3) | 10(21.7) | 0.53 |
| Join and Extended Family | 234(83.6) | 173(73.9) | 61(26.1) |  |
| **Monthly Family income (considering \$1=100 Taka)** | | | | |
| < 10000 Taka (\$100) | 54(19.3) | 38(70.4) | 16(29.6) | 0.701 |
| 10001-20000 Taka (\$100-\$200) | 131(46.8) | 99(75.6) | 32(24.2) | |
| 20001-30000 Taka (\$200-\$300) | 46(16.4) | 33(71.7) | 13(28.3) | |
| > 30000 Taka (>\$300) | 49(17.5) | 39(79.6) | 10(20.4) | |
| **Tobacco user (smoked or smokeless)** |  |  |  |  |
| Yes | 167(59.6) | 131(78.4) | 36(21.6) | 0.05** |
| No | 113(40.4) | 78(69.0) | 35(31.0) |  |
| **Alcohol Consumption** |  |  |  |  |
| Yes | 14(5.0) | 6(42.9) | 8(57.1) | 0.01*** |
| No | 265(94.6) | 202(76.2) | 63(23.8) |  |
| **Daily meal consumption** |  |  |  |  |
| Less than 3 times | 16(5.7) | 11(68.8) | 5(31.3) | 0.153 |
| Three times | 254(90.7) | 193(76.0) | 61(24.0) |  |
| More than three times | 10(3.6) | 5(50.0) | 5(50.0) |  |
| **Meat consumption** |  |  |  |  |
| No | 51(18.2) | 35(68.6) | 16(31.4) | 0.27 |
| Yes | 229(81.8) | 174(76.0) | 55(24.1) |  |
| **Fish consumption** |  |  |  |  |
| No | 15(5.4) | 7(46.7) | 8(53.3) | 0.027** |
| Yes | 265(94.6) | 202(76.2) | 63(23.80) |  |
| **Soft Drink user** |  |  |  |  |
| No | 251(89.6) | 189(75.3) | 62(24.7) | 0.458 |
| Yes | 29(10.4) | 20(69.0) | 9(31.0) |  |
| **Tea Drinker** |  |  |  |  |
| No | 129(46.1) | 103(79.8) | 26(20.2) | 0.05** |
| Yes | 151(53.9) | 106(70.2) | 45(29.8) |  |
| **Sweet eater** |  |  |  |  |
| No | 244(87.1) | 190(77.9) | 54(22.1) | 0.001*** |
| Yes | 36(12.9) | 19(52.8) | 17(47.2) |  |
| #Chi-square/Fisher’s exact test. *Significant at 10% level; **Significant at 5% level; ***Significant at 1% level |  |  |  |  |

The study found that smoking is a common personal habit among tribal elderly and 5% of them were alcohol consumer. Nine out of 10 tribal older people receives three-time meal in a day. However, meat, fish consumption and tea drinking were also found frequent in the community elderly (Table 1).

### Factors associated with Poor oral health status of tribal elderly

Numerous socio-demographic, food consumption behavior and personal habits related factors were responsible with the oral health status of tribal elderly obtained by Pearson’s Chi-square association test (Table 1). Respondents from the higher age group and economically dependent were comparatively experienced to poor oral health status (High DMFT). Variables including marital status, occupation, tobacco and alcohol consumption history, fish and sweet consumption and tea drinker were significantly (p<0.05) associated with the oral health status of tribal elderly.

### Oral health status of the tribal elderly

Represents’ oral health status was assessed by analyzing DMFT index (Fig 1.a, 1.b and Fig 1.c). From the Fig 1.a, it was seen that the prevalence of high DMFT as well as poor oral health status was 25.35%. Which means that one out of four tribal elderly is suffering from poor oral health condition. Fig 1.b shows the elderly oral health status by their age. Analysis found that poor oral health status is age neural and the majority elderly in higher age group are facing poor oral health condition. In addition, poor oral health condition was found vulnerable in female elderly than male (Fig 1.c).

**Fig 1.**
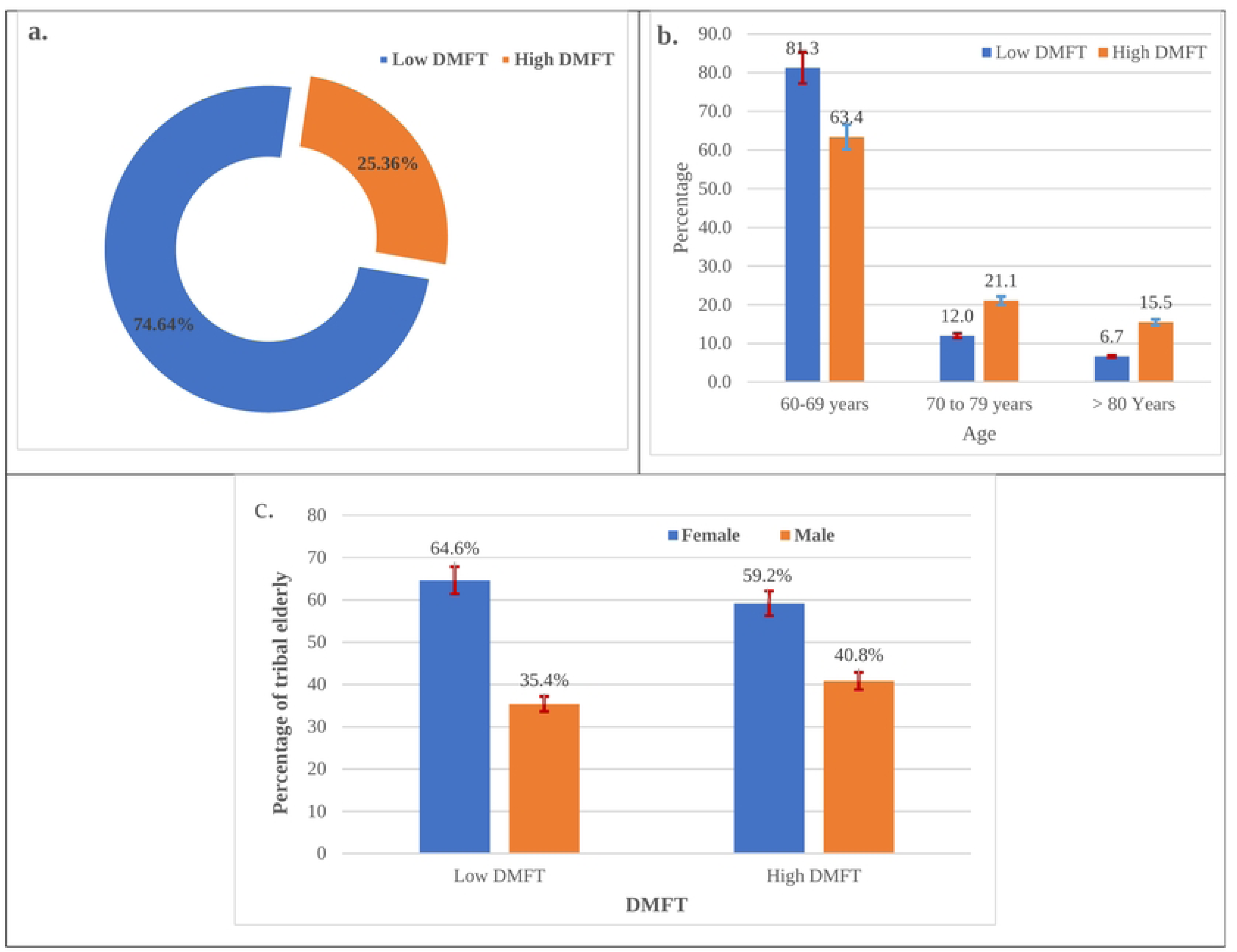
DMFT levels as a predictor of oral health - a. Overall DMFT level; b. DMFT level by age groups and; c. DMFT levels by sex of the tribal elderly in Bangladesh.

### Clinical sign and symptoms related to oral health of the tribal elderly

Respondent’s clinical sign and symptoms related to oral health were analyzed in table 2. It was seen that elderlies were suffering from tooth pain where the problem is high in female than male. Almost all the clinical sign and symptoms including gingival bleeding, dental caries, tooth sensitivity, loose and broken tooth and periodontal pocket presented in Table 2 were found high in female than their male counterparts whereas gingival bleeding was present high among male tribal elderly (Table 2).

**Table 2.** Distribution of clinical sign and symptoms of oral health of the tribal elderly.

| Oral health problems | Male<br>n (%) | Female<br>n (%) | Oral health problems | Male<br>n (%) | Female<br>n (%) |
| --- | --- | --- | --- | --- | --- |
| **Presence of Tooth Pain** |  |  | **Presence of staining in tooth** |  |  |
| Yes | 8(7.9) | 17(9.6) | Yes | 61(35.1) | 113(64.9) |
| No | 93(92.1) | 160(90.4) | No | 42(39.6) | 64(60.4) |
| **Presence of Gingival bleeding** |  |  | **Presence of pain in tongue** |  |  |
| Yes | 9(8.7) | 14(7.9) | Yes | 61(35.1) | 113(64.9) |
| No | 94(91.3) | 163(92.1) | No | 42(39.6) | 64(60.4) |
| **Presence of Dental caries** |  |  | **Presence of lip pain** |  |  |
| Yes | 20(19.4) | 41(23.2) | Yes | 0(0.0) | 1(100) |
| No | 83(80.6) | 136(76.8) | No | 103(36.9) | 176(63.1) |
| **Presence of Sensitivity in tooth** |  |  | **Presence of bad breath** |  |  |
| Yes | 18(32.7) | 37(67.3) | Yes | 20(40.0) | 30(60.0) |
| No | 85(37.8) | 140(62.2) | No | 83(36.1) | 147(63.9) |
| **Presence of loose teeth** |  |  | **Pocket of periodontium by scoring** |  |  |
| Yes | 13(26.5) | 36(73.5) | Presence of Condition (Pocket 4-5 mm) | 6(35.3) | 11(64.7) |
| No | 90(39.0) | 141(61.0) | Absence of Condition | 97(36.9) | 166(63.1) |
| **Presence of Broken tooth** |  |  | **Materials used for oral hygiene practice during toothbrush** |  |  |
| Yes | 29(34.1) | 56(65.9) | Tooth Paste | 64(36.8) | 110(63.2) |
| No | 74(37.9) | 121(62.1) | No Materials used | 2(100) | 0(0.0) |
| **Presence of calculus in tooth** |  |  | Ash | 0(0.0) | 1(100) |
| Yes | 35(35.0) | 65(65) |  |  |  |
| No | 68(37.8) | 112(62.2) |  |  |  |

### Degree of associated factors of oral health status

Numerous socio-demographic, food consumption behavior and personal habits related factors significantly affects the oral health status of tribal elderly found by examining adjusted odds ratio in multivariable logistic regression (Table 3). The chance of affecting to poor oral health condition was comparatively higher in older age groups. More specifically, elderly aged 70 -79 years were 2.46 [AOR: 2.46; 95% confidence Interval (CI): 1.04-5.85] times more and elderly aged 80 or more were 3.33 [AOR: 3.33; 95% confidence Interval (CI): 1.18-9.39] times more likely to suffer poor oral health condition than the young elderly (60 to 69 years). Male elderly was 0.46 [AOR: 0.36; 95% confidence Interval (CI): 0.21-1.01] times lower chance of suffering from poor oral health condition than their female counterparts. Similarly, unmarried and minor religious groups were more likely to suffer from poor oral health condition. Tobacco user were 2.03 [AOR: 2.03; 95% CI: 1.02-4.02] times and alcohol consumers were 6.83 [AOR: 6.83; 95% CI: 1.82-25.62] times more likely to suffer from poor oral health condition than those who did not consumed. Analysis also found that elderly who taken more than three times meal in a day were 3.0 [AOR: 3.0; 95% CI: 0.43-20.72] times and elderly consumed sweet daily were 3.93 [AOR: 3.93; 95% CI: 1.66-9.34] times more likely to affect in poor oral health condition (Table 3).

**Table 3.**
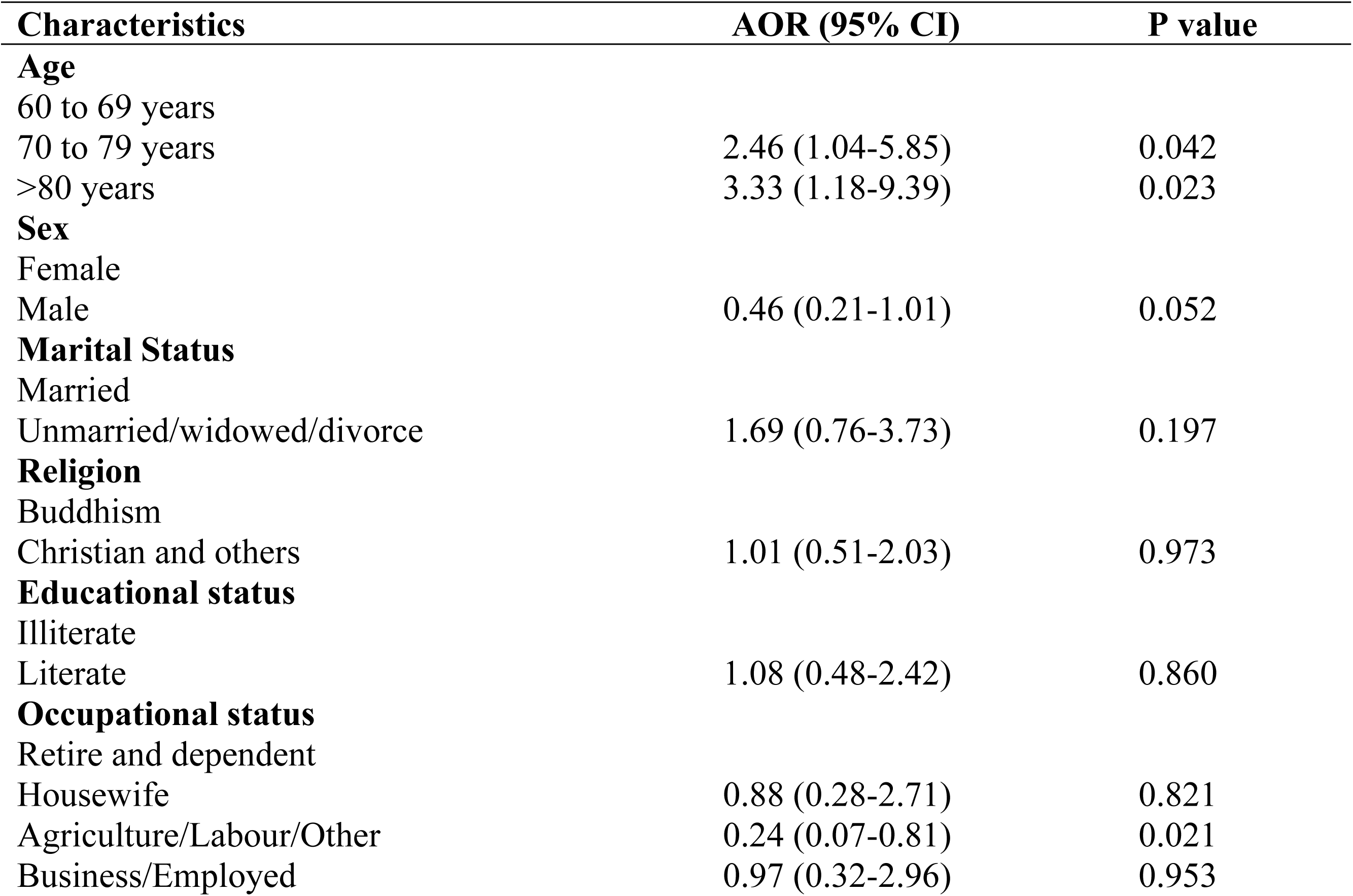
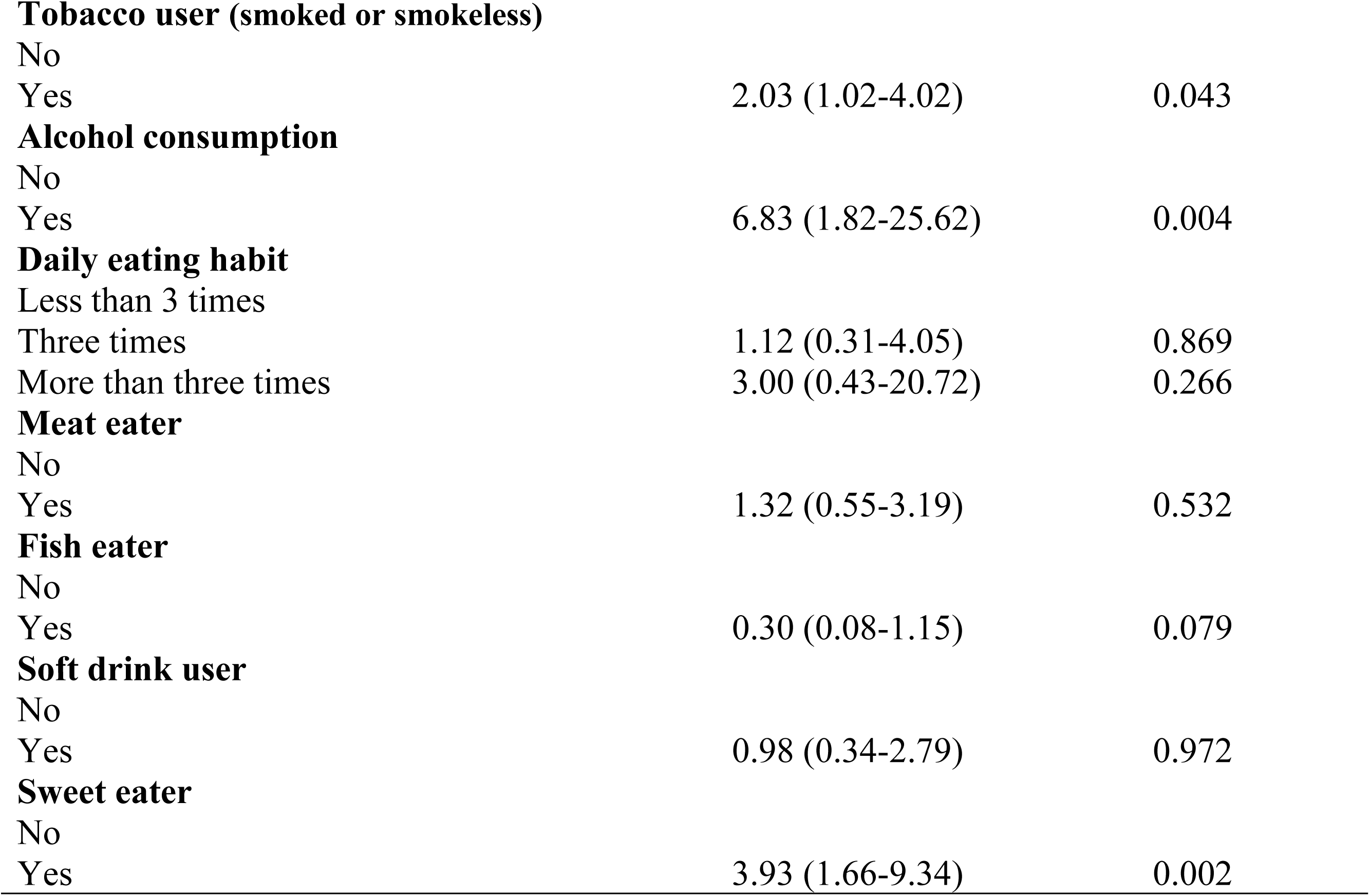
Summary of multivariable logistic regression model of DMFT to assess of oral health status.

| Characteristics | AOR (95% CI) | P value |
| --- | --- | --- |
| **Age** |  |  |
| 60 to 69 years |  |  |
| 70 to 79 years | 2.46 (1.04-5.85) | 0.042 |
| >80 years | 3.33 (1.18-9.39) | 0.023 |
| **Sex** |  |  |
| Female |  |  |
| Male | 0.46 (0.21-1.01) | 0.052 |
| **Marital Status** |  |  |
| Married |  |  |
| Unmarried/widowed/divorce | 1.69 (0.76-3.73) | 0.197 |
| **Religion** |  |  |
| Buddhism |  |  |
| Christian and others | 1.01 (0.51-2.03) | 0.973 |
| **Educational status** |  |  |
| Illiterate |  |  |
| Literate | 1.08 (0.48-2.42) | 0.860 |
| **Occupational status** |  |  |
| Retire and dependent |  |  |
| Housewife | 0.88 (0.28-2.71) | 0.821 |
| Agriculture/Labour/Other | 0.24 (0.07-0.81) | 0.021 |
| Business/Employed | 0.97 (0.32-2.96) | 0.953 |
| **Tobacco user (smoked or smokeless)** |  |  |
| No |  |  |
| Yes | 2.03 (1.02-4.02) | 0.043 |
| **Alcohol consumption** |  |  |
| No |  |  |
| Yes | 6.83 (1.82-25.62) | 0.004 |
| **Daily eating habit** |  |  |
| Less than 3 times |  |  |
| Three times | 1.12 (0.31-4.05) | 0.869 |
| More than three times | 3.00 (0.43-20.72) | 0.266 |
| **Meat eater** |  |  |
| No |  |  |
| Yes | 1.32 (0.55-3.19) | 0.532 |
| **Fish eater** |  |  |
| No |  |  |
| Yes | 0.30 (0.08-1.15) | 0.079 |
| **Soft drink user** |  |  |
| No |  |  |
| Yes | 0.98 (0.34-2.79) | 0.972 |
| **Sweet eater** |  |  |
| No |  |  |
| Yes | 3.93 (1.66-9.34) | 0.002 |

## Discussion

Research on the oral health of tribal elderly is rarely conducted in Bangladesh. Although, the government has some special health policy focus on overall or main-land elderly but tribal elderly in Bangladesh is still vulnerable due to their geographic location. This pilot study cross-sectionally collected and analyzed oral health related data of 280 tribal elderly residing Bandarban-a main tribal residing region in Bangladesh. The socio-demographic and economic condition of the tribal elderly was poor. The female elderly and Buddhist religion were comparatively higher than their counterparts.

In Bangladesh, more than one out of five tribal elderlies were suffering from poor oral health status. The rate was very high among female and higher age groups. Generally, elderly from higher age group and females are reluctant to maintain oral health due to their frail physical condition [43,44]. As a results, they comparatively suffer more in poor oral health condition. This study revealed that sociodemographic traits, personal habits like alcohol and cigarette use, dietary habits like eating meat and fish for every meal, drinking soft drinks, drinking tea, and eating sweets, all are linked to poor oral health condition (High DMFT). According to the World Health Organization (WHO), elderlies’ poor oral health is caused by some modifiable risk factors including tea and sugar consumption, tobacco use, alcohol use, poor hygiene practice, and their underlying socio-demographic and commercial determinants [45]. In contrast, K Bruno-Ambrosius’s study conducted among Swedish female teenagers showed that tooth brushing practices, sweets, and snacks did not significantly affect the development of caries in Swedish female teenagers [46].

The results of this study indicate that the oral health status of the tribal elderly population was poor, with a prevalence of 25.35% of high DMFT, indicating poor oral health. This suggests that one out of four tribal elderly individuals are suffering from poor oral health. A recent health care study supports this finding by demonstrating the oral health issues that tribal groups faced [47]. Numerous literatures suggests that poor oral health status (high DMFT) is negatively associated with aging and it affects the oral health related quality of life of elderly aged 75 years and more [48,49]. The results of this research suggest that elderly individuals are more likely to suffer from poor oral health condition than younger individuals. Specifically, those aged 70-79 years were 2.46 times more likely and those aged 80 or more were 3.33 times more likely to suffer from poor oral health condition than the young elderly (60 to 69 years). Additionally, male elderly were 0.46 times less likely to suffer from poor oral health condition than their female counterparts.

Unmarried and minor religious groups were also more likely to suffer from poor oral health condition. The finding is consistent with previous research that has found that older adults are at an increased risk for poor oral health and that women tend to have poorer oral health than men [48,50]. Furthermore, tobacco users were 2.03 times more likely and alcohol consumers were 6.83 times more likely to suffer from poor oral health condition than those who did not consume either substance. Lastly, elderly who took more than three meals a day were 3.0 times more likely and those who consumed sweets daily were 3.93 times more likely to suffer from poor oral health condition. In line with these findings, it is evident that sugary diets, tobacco use, and alcohol intake posed significant risks for dental illnesses, discomfort, and function impairment reported the World Health Organization [19].

Most of the tribal elderly in Bangladesh live in hilly and hard-to-reach area. Due to the challenging terrain and limited infrastructure, there is a limited access to healthcare services, including dental care. However, improving the oral health vulnerability among tribal elderly requires a comprehensive approach-

**Increasing access to dental care-** Mobile dental clinics and outreach programs should be established in remote tribal areas to provide check-ups, treatments, and oral health education. Collaborations with local healthcare organizations and dental professionals can ensure affordable services in areas with limited healthcare facilities.

**Raising oral health awareness-** Tailored education programs should emphasize proper oral hygiene practices, like regular brushing and flossing. Training programs for community health workers can enable oral health assessments and promote preventive measures [38]. Partnerships with dental professionals can establish referral systems for specialized care.

**Collaboration among stakeholders-** Government agencies, NGOs, dental associations, and tribal leaders should develop comprehensive oral health programs. International organizations and donor agencies can provide funding support.

**Research and data collection-** Studies should assess oral health status and needs among tribal elderly in different regions. Data on oral health indicators will facilitate monitoring and evaluation.

**Policy development**-Advocacy for policies that allocate resources and establish guidelines for tribal oral healthcare is necessary. Integrating oral health within existing healthcare policies will ensure holistic care.

## Conclusion

In conclusion, this study has highlighted the poor oral health status of tribal elderly in Bangladesh. The findings suggest that elderly individuals are more likely to suffer from poor oral health condition than younger individuals, and that certain lifestyle factors such as tobacco and alcohol consumption, as well as meal frequency and sweet consumption, can further increase the risk of poor oral health condition in the elderly. It is important to note that these findings should be taken into consideration when developing strategies to improve oral health among the elderly population.

## Acknowledgement

National institute of Preventive and Social Medicine (NIPSOM), Mohakhali, Dhaka-1212 is an apex body for public health education and research in Bangladesh. We thank the Department of Biostatistics, NIPSOM for their technical support during the study. We are thankful to the participants who provided time and shared their health experiences.

## Ethics approval statement

This study followed the guidelines of World Medical Association (WMA) Declaration of Helsinki. The ethical approval of this study was approved by the institutional review board of National Institute of Preventive and Social Medicine (NIPSPOM), Mohakhali, Dhaka, Bangladesh under the registration number NIPSOM/IRB/2019/111. The formal permission of data collection in the community was taken from the tribal community leaders (called ‘Karbari’). Both written and verbal consent were taken from the participants before initiating the interview.

## Patient consent statement

Both written and verbal consent was taken from each participant before initiating the interview for data collection. A brief introduction on the aims and objectives of the study was given first and then, the written consent translated in native language was read out for illiterate tribal elderly. Participants who were agreed with the consent were finally included in the study.

## Author’s contributions

**Shomrita Barua**- Contributed in Conception, data collection, analysis, writing original draft, editing and finalizing the version to be submitted. **Sunanda Bosu, Mohammad Farhadul Haque & Liza Bosak**- Data collection, writing original draft, editing and finalizing the version to be submitted. **Md Rezowan Rashid, Shaikh Riaduz Zaman & Md. Foyzur Rahman**- Writing original draft, editing and finalizing the version to be submitted. **Mohammad Meshbahur Rahman**- Contributed in conception of the study, supervised, analyzed, writing original draft, editing and finalizing the version to be submitted. All author has read and approved the final version of the manuscript to be submitted.

## Data availability statement

The datasets generated and/or analyzed during the current study are available from the corresponding author on reasonable request to.

## References

1. Chan AKY, Tamrakar M, Jiang CM, Lo ECM, Leung KCM, Chu C-H. Common Medical and Dental Problems of Older Adults: A Narrative Review. Geriatrics (Basel). 2021;6. doi:10.3390/geriatrics6030076

2. Griffin SO, Jones JA, Brunson D, Griffin PM, Bailey WD. Burden of oral disease among older adults and implications for public health priorities. Am J Public Health. 2012;102: 411–418. doi:10.2105/AJPH.2011.300362

3. Nicksic NE, Massie AW, Byrd-Williams CE, Kelder SH, Sharma S V, Butte NF, et al. Dietary Intake, Attitudes toward Healthy Food, and Dental Pain in Low-Income Youth. JDR Clin Trans Res. 2018;3: 279–287. doi:10.1177/2380084418774039

4. Azzolino D, Passarelli PC, De Angelis P, Piccirillo GB, D’Addona A, Cesari M. Poor Oral Health as a Determinant of Malnutrition and Sarcopenia. Nutrients. 2019;11. doi:10.3390/nu11122898

5. Burt BA, Eklund SA, Morgan KJ, Larkin FE, Guire KE, Brown LO, et al. The Effects of Sugars Intake and Frequency of Ingestion on Dental Caries Increment in a Three-year Longitudinal Study. J Dent Res. 1988;67: 1422–1429. doi:10.1177/00220345880670111201

6. Millen AE, Dahhan R, Freudenheim JL, Hovey KM, Li L, McSkimming DI, et al. Dietary carbohydrate intake is associated with the subgingival plaque oral microbiome abundance and diversity in a cohort of postmenopausal women. Sci Rep. 2022;12: 2643. doi:10.1038/s41598-022-06421-2

7. Rooban T, Vidya KM, Joshua E, Rao A, Ranganathan S, Rao UK, et al. Tooth decay in alcohol and tobacco abusers. Journal of Oral and Maxillofacial Pathology. 2011;15: 14–21. doi:10.4103/0973-029X.80032

8. Dasanayake AP, Warnakulasuriya S, Harris CK, Cooper DJ, Peters TJ, Gelbier S. Tooth Decay in Alcohol Abusers Compared to Alcohol and Drug Abusers. Vieira AR, editor. Int J Dent. 2010;2010: 786503. doi:10.1155/2010/786503

9. Lula ECO, Ribeiro CCC, Hugo FN, Alves CMC, Silva AAM. Added sugars and periodontal disease in young adults: an analysis of NHANES III data. Am J Clin Nutr. 2014;100: 1182–1187. doi:10.3945/ajcn.114.089656

10. Martini D, Galli C, Guareschi C, Angelino D, Bedogni G, Biasini B, et al. Claimed effects, outcome variables and methods of measurement for health claims on foods proposed under Regulation (EC) 1924/2006 in the area of oral health. NFS Journal. 2018;10: 10–25. doi:https://doi.org/10.1016/j.nfs.2017.12.001

11. Chen H, Zhang R, Cheng R, Xu T, Zhang T, Hong X, et al. Gingival bleeding and calculus among 12-year-old Chinese adolescents: a multilevel analysis. BMC Oral Health. 2020;20: 147. doi:10.1186/s12903-020-01125-3

12. Rahman M, Hamiduzzaman M, Akter M, Farhana Z, Hossain M, Hasan M, et al. Frailty indexed classification of Bangladeshi older adults’ physio-psychosocial health and associated risk factors-a cross-sectional survey study. BMC Geriatr. 2021;21: 3. doi:10.1186/s12877-020-01970-5

13. Paul GK, Rahman MM, Hamiduzzaman M, Farhana Z, Mondal SK, Akter S, et al. Hypertension and its physio-psychosocial risks factors in elderly people: a cross-sectional study in north-eastern region of Bangladesh. Journal of geriatric cardiology. 2021;18: 75–82. doi:10.11909/j.issn.1671-5411.2021.01.011

14. Rahman M, Begum M, Uddin M, Rahman M. Factors Affecting Health Status of Urban Aged Population: Evidence from Sylhet, Bangladesh. Indian Journal of Gerontology. 2018;32: 103–118.

15. Bassim CW, MacEntee MI, Nazmul S, Bedard C, Liu S, Ma J, et al. Self-reported oral health at baseline of the Canadian Longitudinal Study on Aging. Community Dent Oral Epidemiol. 2020;48: 72–80. doi:10.1111/cdoe.12506

16. Calzada MT, Posada-López A, Gutiérrez-Quiceno B, Botero JE. Association Between Tobacco Smoking, Dental Status and Self-perceived Oral Health in Elderly Adults in Colombia. J Cross Cult Gerontol. 2021;36: 187–200. doi:10.1007/s10823-021-09426-y

17. Ghosal S, Sinha A, Kerketta S, Acharya AS, Kanungo S, Pati S. Oral health among adults aged ≥45 years in India: Exploring prevalence, correlates and patterns of oral morbidity from LASI wave-1. Clin Epidemiol Glob Health. 2022;18: 101177. doi:https://doi.org/10.1016/j.cegh.2022.101177

18. Neelamana SK, Janakiram C, Varma B. Oral health status and related quality of life among elderly tribes in India. J Family Med Prim Care. 2020;9. Available: https://journals.lww.com/jfmpc/Fulltext/2020/09120/Oral_health_status_and_related_quality_of_life.28.aspx

19. Petersen PE. The World Oral Health Report 2003: continuous improvement of oral health in the 21st century – the approach of the WHO Global Oral Health Programme. Community Dent Oral Epidemiol. 2003;31: 3–24. doi:https://doi.org/10.1046/j..2003.com122.x

20. Lukacs JR. Gender differences in oral health in South Asia: Metadata imply multifactorial biological and cultural causes. American Journal of Human Biology. 2011;23: 398–411. doi:https://doi.org/10.1002/ajhb.21164

21. World Health Organization. Action plan for oral health in South-East Asia 2022–2030. 25 Oct 2022 [cited 28 Jan 2023]. Available: https://www.who.int/publications/i/item/9789290210061

22. Prasai Dixit L, Shakya A, Shrestha M, Shrestha A. Dental caries prevalence, oral health knowledge and practice among indigenous Chepang school children of Nepal. BMC Oral Health. 2013;13: 20. doi:10.1186/1472-6831-13-20

23. Singh A, Purohit BM. Addressing oral health disparities, inequity in access and workforce issues in a developing country. Int Dent J. 2013;63: 225–229. doi:https://doi.org/10.1111/idj.12035

24. Roy P, Deshwara M. Ethnic population in 2022 census: Real picture not reflected. The Daily Star. 7 Jan 2023.

25. Rahman SA, Kielmann T, McPake B, Normand C. Healthcare-seeking Behaviour among the Tribal People of Bangladesh: Can the Current Health System Really Meet Their Needs? J Health Popul Nutr. 2012;30: 353–365. doi:https://doi.org/10.3329/jhpn.v30i3.12299

26. Jabeen S, Manni U, Shakil S. Oral health status among tobacco users in the selected rural population. Bangladesh Med J. 2014;43. doi:https://doi.org/10.3329/bmj.v43i2.21387

27. Zhang Yixin and He J and HB and HR and LM. Effect of tobacco on periodontal disease and oral cancer. Tob Induc Dis. 2019;17. doi:10.18332/tid/106187

28. Heck JE, Marcotte EL, Argos M, Parvez F, Ahmed A, Islam T, et al. Betel quid chewing in rural Bangladesh: prevalence, predictors and relationship to blood pressure. Int J Epidemiol. 2012;41: 462–471. doi:10.1093/ije/dyr191

29. Sa S, Haseen F, Ss I, Sf C. Knowledge and Practice of Oral Health and Hygiene and Oral Health Status among School Going Adolescents in a Rural Area of Sylhet District, Bangladesh. CBMJ. 2021.

30. Kabir MN, Ahmed MB, Khan M. Knowledge and Oral Hygiene Practice by School Children in Cox’s Bazar, Bangladesh. Update Dental College Journal. 2019;9: 27–31. doi:10.3329/updcj.v9i2.43736

31. Sayeed Ahmad M, Abdullah Al-Mamun M, Begum S, Shahidul Islam M, Ahsan Habib M, Mahafuzur Rahman M. Knowledge and Practice About Oral Hygiene by Tribal People (Orao) in Rangpur Region. Bangladesh International Journal of Dental Medicine. 2015;1: 28–32. doi:10.11648/j.ijdm.20150103.12

32. Iqbal MdA, Mohol J, Afrin F, Khaleque MdA, Hohra F-T, Jannat N. Prevalence of periodontal diseases among the patient visiting at Periodontology OPD Update Dental College Hospital, Dhaka. Updat Dent Coll J. 2015;5: 23–29. doi:https://doi.org/10.3329/updcj.v9i2.43733

33. van Palenstein Helderman WH, Joarder MA, Begum A. Prevalence and severity of periodontal diseases and dental caries in Bangladesh. Int Dent J. 1996;46: 76–81. Available: http://europepmc.org/abstract/MED/8930677

34. Pearson N, Croucher R, Marcenes W, O’Farrell M. Dental health and treatment needs among a sample of Bangladeshi medical users aged 40 years and over living in Tower Hamlets, UK. Int Dent J. 2001;51: 23–29. doi:https://doi.org/10.1002/j.1875-595X.2001.tb00813.x

35. Mth C, Mai K, Afma C. Prevalence of Gingivitis, Plaque accumulation and Decayed, Missing and Filled Teeth among slum population in Bangladesh. Bangladesh Med Res Counc Bull. 2014.

36. World Health Organization. Global oral health status report: towards universal health coverage for oral health by 2030. 2022 Nov. Available: https://www.who.int/publications/i/item/9789240061484

37. Davy C, Harfield S, McArthur A, Munn Z, Brown A. Access to primary health care services for Indigenous peoples: A framework synthesis. Int J Equity Health. 2016;15: 163. doi:10.1186/s12939-016-0450-5

38. Chowdhury S, Roy S, Hasan M, Sadique A Al, Islam T, Hasan M, et al. Oral health knowledge, practice, and oral health status among rohingya refugees in Cox’s Bazar, Bangladesh: A cross-sectional study. PLoS One. 2022;17: e0269359-. Available: https://doi.org/10.1371/journal.pone.0269359

39. Ahmad M, Al-Mamun M, Islam M, Siddik A, Rahman M, Asaduzzaman H. Oral hygiene practice and oral health status of geriatric population in selected area of Rangpur, Bangladesh. KYAMC Journal. 2018. doi:https://doi.org/10.3329/kyamcj.v9i2.38146

40. Preshaw PM. Detection and diagnosis of periodontal conditions amenable to prevention. BMC Oral Health. 2015;15 Suppl 1: S5. doi:10.1186/1472-6831-15-S1-S5

41. World Health Organization. Oral health surveys : basic methods. 5th ed. WHO, editor. World Health Organization; 2013.

42. Paul G, Rahman MM, Naznin S, Chowdhury M, Uddin MJ. Depression and Anxiety among University Students: A Comparison between COVID-19 Pandemic Panic Period and Post-panic Period in Bangladesh. Open Access Maced J Med Sci. 2022;10: 52–59. doi:10.3889/oamjms.2022.7559

43. Hakeem FF, Bernabé E, Sabbah W. Association between oral health and frailty: A systematic review of longitudinal studies. Gerodontology. 2019;36: 205–215. doi:https://doi.org/10.1111/ger.12406

44. Kim H, Lee E, Lee S-W. Association between oral health and frailty: results from the Korea National Health and Nutrition Examination Survey. BMC Geriatr. 2022;22: 369. doi:10.1186/s12877-022-02968-x

45. World Health Organization. Oral health. In: WHO [Internet]. 18 Nov 2022 [cited 16 Jan 2023]. Available: https://www.who.int/news-room/fact-sheets/detail/oral-health

46. Bruno-Ambrosius K, Swanholm G, Twetman S. Eating habits, smoking and toothbrushing in relation to dental caries: a 3-year study in Swedish female teenagers. Int J Paediatr Dent. 2005;15: 190–196. doi:https://doi.org/10.1111/j.1365-263X.2005.00621.x

47. Cladoosby B (Speepots). Indian Country Leads National Movement to Knock Down Barriers to Oral Health Equity. Am J Public Health. 2017;107: S81–S84. doi:10.2105/AJPH.2017.303663

48. Baniasadi K, Armoon B, Higgs P, Bayat A-H, Mohammadi Gharehghani MA, Hemmat M, et al. The Association of Oral Health Status and socio-economic determinants with Oral Health-Related Quality of Life among the elderly: A systematic review and meta-analysis. Int J Dent Hyg. 2021;19: 153–165. doi:https://doi.org/10.1111/idh.12489

49. Patel J, Wallace J, Doshi M, Gadanya M, ben Yahya I, Roseman J, et al. Oral health for healthy ageing. Lancet Healthy Longev. 2021;2: e521–e527. doi:https://doi.org/10.1016/S2666-7568(21)00142-2

50. Ferraro M, Vieira AR. Explaining Gender Differences in Caries: A Multifactorial Approach to a Multifactorial Disease. Seymen F, editor. Int J Dent. 2010;2010: 649643. doi:10.1155/2010/649643

